# Systematic Review and Meta-Analysis of Stem Cell Therapy in Myocardial Infarction: Effects on Left Ventricular Ejection Fraction and Major Adverse Cardiovascular Events

**DOI:** 10.1101/2025.08.25.25334375

**Authors:** Manish Juneja, Rakhshanda Khan, Mohammed Mussawir Khan, Naga Sai Gouthami Gurujala, Mohd Sijad Uddin, Areej Fatima, Aron Shrey Rebello, Manav Kamleshkumar Patel, Saket Dineshkumar Prajapati, Rohan Singhal, Shrouk Ramadan, Harshawardhan Dhanraj Ramteke

## Abstract

**Introduction:** Acute myocardial infarction (AMI) is a leading cause of morbidity and mortality worldwide, often progressing to heart failure (HF) despite advances in treatment. Stem cell therapy has emerged as a promising approach to repair myocardial tissue, improve cardiac function, and reduce the progression to HF. However, the long-term efficacy and clinical outcomes remain uncertain. This systematic review and meta-analysis aimed to evaluate the mid-to long-term effects of stem cell therapy on left ventricular ejection fraction (LVEF), infarct size, and major adverse cardiovascular events (MACE) in AMI patients.

**Methods:** A comprehensive search was conducted across PubMed, Scopus, Embase, and CENTRAL databases, up to August 2025, following PRISMA guidelines. The study included randomized controlled trials (RCTs) that assessed the effects of stem cell therapy on LVEF and MACE. Data extraction was performed independently by two reviewers, and risk of bias was assessed using Cochrane’s Risk of Bias 2.0 tool. Statistical analyses were performed using Stata 18.0, with heterogeneity quantified using I², and publication bias assessed through funnel plots.

**Results:** A total of 83 studies involving 7307 patients were included in the analysis. The average age of the patients was 55.29 ± 12 years, with a follow-up duration of 27.5 days. Significant improvements in LVEF were observed at various time points: 1.83% at 6 months, 2.21% at 12 months, and 2.21% at 24 months. The highest mean difference in LVEF was observed in Wang et al. (2014) with a mean difference (MD) of 17.60%, while the lowest was −4.20% in Wohrle et al. (2010). MACE analysis showed a favorable trend at 6 months with an odds ratio of −0.89 (95% CI: −1.14; −0.63), but no significant improvement at 12, 24, or 36 months. A reduction in infarct size was noted at 6 months (MD: 1.80%, 95% CI: 0.58; 3.02), but this was minimal at 12 months (MD: 0.70%, 95% CI: 0.48; 0.91).

**Conclusion:** Stem cell therapy shows promise for improving LVEF and reducing infarct size in the short to mid-term after AMI. However, long-term benefits remain inconclusive, with minimal effects observed at 24 and 36 months. Further studies are required to determine the optimal timing, dosage, and stem cell source to maximize therapeutic efficacy. Additionally, the use of major adverse cardiovascular events (MACE) as a primary endpoint may provide a more comprehensive assessment of the long-term impact of stem cell therapy in AMI patients.

## 1. Introduction

Acute myocardial infarction (AMI) remains one of the leading causes of morbidity and mortality worldwide, despite substantial advances in diagnostic tools, treatment strategies, and patient management in the past decades. AMI is a primary precipitant of heart failure (HF), a progressive condition that dramatically impairs quality of life and contributes significantly to the growing burden on healthcare systems globally [1]. The prevalence of heart failure in patients who survive AMI is increasing, with approximately 10-15% of individuals who experience AMI developing chronic heart failure [2]. This progression to HF is linked with poor clinical outcomes, including repeated hospitalizations, diminished life expectancy, and high treatment costs [3]. Thus, strategies aimed at preserving or improving left ventricular function in the aftermath of AMI are critical to mitigating these outcomes and enhancing survival.

The conventional management of AMI focuses primarily on early reperfusion, which aims to restore blood flow and limit myocardial injury. However, despite the remarkable advances in pharmacological therapies and mechanical interventions such as percutaneous coronary intervention (PCI), the resulting cardiac dysfunction often remains irreversible, leading to long-term complications [4]. This underscores the need for innovative approaches that not only stabilize patients in the short term but also prevent or reverse the long-term damage caused by AMI. Among such therapies, stem cell-based regenerative strategies have garnered significant attention due to their potential to repair or regenerate damaged myocardial tissue, restore cardiac function, and reduce the progression to heart failure.

Stem cell therapy offers considerable promise for myocardial repair following AMI. The potential regenerative ability of stem cells to repair tissue damage and promote functional recovery stems from their capacity to differentiate into myocardial cells, secrete paracrine factors that enhance tissue regeneration, and support angiogenesis [5]. Numerous studies have explored various types of stem cells, including mesenchymal stem cells (MSCs), induced pluripotent stem cells (iPSCs), and cardiac progenitor cells (CPCs), in animal models and clinical trials. Early studies have shown that stem cell transplantation can improve left ventricular ejection fraction (LVEF), reduce infarct size, and improve the overall prognosis of patients with AMI [6]. However, the clinical translation of these promising results has been hindered by inconsistent outcomes, with long-term studies failing to confirm sustained benefits, primarily due to differences in study design, stem cell source, and administration protocols.

One of the primary challenges of stem cell therapy in AMI is determining the optimal timing and cell quantity for effective therapy. While studies have suggested that earlier administration of stem cells may offer superior results in terms of functional recovery, the risks associated with premature cell infusion—such as the potential for arrhythmias and immune rejection—remain significant [7]. Conversely, administering stem cells too late, after significant myocardial damage has already occurred, may limit their therapeutic potential. There is also the issue of adequate stem cell quantity, as the number of stem cells necessary to achieve a meaningful therapeutic effect is still uncertain. Larger quantities of stem cells are often required to generate therapeutic benefits, but harvesting and culturing sufficient cells is a complex and resource-intensive process. Furthermore, while autologous stem cell therapy (using the patient’s own cells) mitigates the risk of immune rejection, it also introduces challenges in cell harvesting, culture time, and handling [8]. Thus, there is a need for further research to determine the optimal cell dose and timing of administration, which would maximize therapeutic efficacy while minimizing risks.

An essential aspect of evaluating the success of stem cell therapy in AMI is the selection of appropriate outcome measures. Traditionally, cardiac function has been assessed through parameters such as LVEF, infarct size, and left ventricular end-diastolic volume (LVEVD), which are frequently measured using imaging techniques such as echocardiography and magnetic resonance imaging (MRI) [9]. These measures, while informative, are often subject to operator variability and can be influenced by various external factors, which makes them less reliable in some clinical settings. Moreover, while improvements in LVEF and infarct size are indicative of myocardial recovery, they do not necessarily translate into improved patient outcomes. As a result, there has been increasing interest in using major adverse cardiovascular events (MACEs) as a more comprehensive outcome measure. MACEs, which include cardiovascular death, reinfarction, stroke, and hospitalization for heart failure, provide a more clinically relevant assessment of the safety and efficacy of treatment strategies [10]. Given the significant impact that AMI-related complications have on patient morbidity and mortality, MACE represents a critical composite endpoint for evaluating the success of stem cell therapy in this patient population.

Despite the progress made in understanding the role of stem cell therapy in AMI, several gaps remain in the current literature. While short-term studies have demonstrated improvements in cardiac function following stem cell administration, the long-term efficacy and safety of stem cell therapy remain inconclusive. There is a need for well-designed, large-scale clinical trials that assess both the immediate and extended benefits of stem cell therapy, taking into account the optimal timing, dose, and type of stem cells used. Additionally, the use of MACEs as a primary endpoint, in combination with more objective measures of cardiac function, may provide a more reliable and clinically relevant assessment of treatment efficacy.

This systematic review aims to critically evaluate the existing evidence on the mid-to long-term effectiveness of stem cell therapy in patients with AMI. Specifically, we will assess the impact of stem cell therapy on LVEF, infarct size, and MACE, with a focus on identifying the optimal cell dose and administration timing that maximize therapeutic benefits while ensuring patient safety. Through this analysis, we hope to provide a clearer understanding of the potential role of stem cell therapy in the management of AMI and contribute to the development of more effective treatment strategies for heart failure prevention and myocardial regeneration.

## 2. Methods

### 2.1 Literature Search

This meta-analysis was conducted following a pre-specified protocol and is reported in accordance with the PRISMA guidelines [11]. A comprehensive search was performed across several databases, including PubMed, Scopus, Embase, and CENTRAL, with the search extended until August 2025. The study protocol is registered with PROSPERO (International Prospective Register of Systematic Reviews) under the registration number The search was designed to identify studies involving patients diagnosed with myocardial infarction (MI) who were treated with stem cell therapy, with standard care or placebo as the comparator, and outcomes focusing on left ventricular ejection fraction (LVEF) as the primary outcome and major adverse cardiovascular events (MACE) as the secondary outcome. Only randomized controlled trials (RCTs) were included to ensure the highest level of evidence. The search strategy was guided by the PICOS framework and used Boolean expressions to ensure thorough identification of relevant studies. Additionally, the reference lists of eligible articles were hand-searched to identify any additional studies that may have been missed in the initial search. No language restrictions were applied. The full search string used in the search process is provided in the Supplementary File.

### 2.2 Screening

All citations retrieved from the four databases were exported into Rayyan, a web-based platform for screening and de-duplication, where duplicates were identified and removed. We then applied a priori eligibility criteria for quantitative synthesis: (i) peer-reviewed, full-text publications in English; (ii) exclusion of case reports, protocols, letters, narrative/systematic reviews and meta-analyses, conference abstracts, ongoing/unpublished studies, and observational designs; (iii) randomized controlled trials with complete outcomes; (iv) intervention arms with Stem Cell treatment and LVEF Changes, infarct size changes and MACE.

### 2.3 Data Extraction and Statistical Analysis

Two reviewers independently extracted data into Microsoft Excel 2021; discrepancies were resolved by consensus. Variables were grouped into three domains: (i) demographics (publication year, first author, sample size, age range); (ii) Acute Myocardial Infarction and (iii) MACE, change in infarct size, change in LVEF were mentioned. When medians and interquartile ranges were reported, values were converted to means and standard deviations using standard methods. Along with that Risk Ratio was presented in forest plots. Heterogeneity was quantified with I²; random-effects models were used for I²>50%, otherwise fixed-effects were applied. Analyses were performed in Stata 18.0. Funnel plots were used for publication bias.

### 2.4 Risk of Bias

The Risk of Bias was analyzed using Cochranes Risk of Bias 2.0 tool and ROBINS-I, which is for Randomized Controlled Trials. Funnel Plots were made for analysis for publication bias, and furthermore, Grade Analysis was done to assess the Quality of Studies.

## 3. RESULTS

### 3.1 Demographics

A total of 1662 records were analyzed, out of which 792 were duplicate records, and 75 were marked as ineligible, out of which 757 were analyzed, and 482 were excluded, hence the finally 275 were extracted for full text screening and 83 were later taken up for final screening, out of which 77 [12–88] were included in study. Total of 7307 patients were analyzed, 5324 were male patients and 1737 were females. The total number of patients in treatment group were 4149 and control group were 3161. The average age of the patients were 55.29 ± 12 years, the average duration of follow up was 27.5 days and duration of culture was 2.05 ± 0.65 week. The follow up time was 16.01 ± 7 days. The total number of patients receiving Allogeneic Cardiac Stem Cells (AlloCSC-01) were 33, Bone Marrow Mesenchymal Stem Cells (BMMSCs) were 3562, BPMCs (Bone Marrow Peripehral Mononuclear Cells) were 33, CD 133+ were 20, CD 34+ were 53, G-CSF were 301, Human Mesenchymal Stem Cells (hMSCs) were 39, Mesenchymal Stem Cells (MSCs) were 10, PBMNCs (Peripheral Blood Mononuclear Cells) were 10, WJ-MSCs (Whartons Jelly Mesenchymal Stem Cells) were 58.

### 3.2 Change in LVEF at Baseline

A total of 73 studies were analyzed, which showed pooled Mean difference of LVEF of 0.22 (95% CI: −0.48; 0.91). The Highest mean difference was seen in Wang et. Al. 2014, with Mean Difference (MD) of 17.60 (95% CI: 15.16; 20.04). The lowest mean difference was seen in Wohrle et. Al. 2010, with MD −4.20 (95% CI: −8.38; −0.02). The Heterogeneity was 6.93 and I-Square was 96.49, making the results not significant. The forest plot is in Figure 2.

### 3.3 Change in LVEF at 6 months

A total of 63 studies were analyzed, which showed pooled Mean difference of LVEF of 1.83 (95% CI: 0.98; 2.69). The Highest mean difference was seen in Lezo et. Al. 2007, with Mean Difference (MD) of 13.00 (95% CI: 6.72; 19.28). The lowest mean difference was seen in Wohrle et. Al. 2010, with MD −7.5 (95% CI: −10.69; - 4.31). The Heterogeneity was 9.49 and I-Square was 96.83, making the results not significant. The forest plot is in Figure 3.

### 3.4 Change in LVEF at 12 months

A total of 24 studies were analyzed, which showed pooled Mean difference of LVEF of 2.21 (95% CI: 0.94; 3.47). The Highest mean difference was seen in Yang et. Al. 2020, with Mean Difference (MD) of 9.50 (95% CI: 5.20; 13.80). The lowest mean difference was seen in Wohrle et. Al. 2010, with MD −5.20 (95% CI: −11.85; 1.45). The Heterogeneity was 7.43 and I-Square was 97.01, making the results not significant. The forest plot is in Figure 4.

### 3.5 Change in LVEF at 24 Months

A total of 9 studies were analyzed, which showed pooled Mean difference of LVEF of 1.57 (95% CI: - 0.65,3.79). The Highest mean difference was seen in Assmus et. Al. 2010, with Mean Difference (MD) of 6.50 (95% CI: 6.03; 6.97). The lowest mean difference was seen in Wohrle et. Al. 2010, with MD −6.60 (95% CI: - 12.97; −0.23). The Heterogeneity was 8.37 and I-Square was 96.36, making the results not significant. The forest plot is in Figure 5.

### 3.6 Change in LVEF in 36 months

A total of 5 studies were analyzed, which showed pooled Mean difference of LVEF of 1.46 (95% CI: - 0.39,3.32). The Highest mean difference was seen in Cao et. Al. 2009, with Mean Difference (MD) of 4.10 (95% CI: 3.76; 4.44). The lowest mean difference was seen in Meyer et. Al. 2009, with MD -6.60 (95% CI: -8.15; 6.95). The Heterogeneity was 3.57 and I-Square was 94.46, making the results not significant. The forest plot is in Figure 6.

### 3.7 MACE interpretation at 6 Months

A total of 23 studies were analyzed, which showed pooled Odds ratio of MACE was -0.89 (95% CI: -1.14;-0.63). The highest odds of MACE were Wohrle et. Al. 2013 0.92 (95% CI: -1.34; 3.17). The lowest odds ratio was seen in Tendera et. Al. 2009 -2.07 (95% CI: -2.87; 0.52). The Heterogeneity was 1.15 and I-Squared was 13.18. The Forest Plot in Figure 7.

### 3.8 MACE interpretation at 12 Months

A total of 13 studies were analyzed, which showed odds ratio of MACE was -0.62 (95% CI: -0.90; -0.35). The Highest odds of MACE were in -0.88 (95% CI: -1.84; 0.07). The lowest odds ratio was -2.05 (95% CI: -3.13; - 0.98). The Heterogeneity was 20.26 and I-Squared was 1.25. The Forest Plot in Figure 8.

### 3.9 MACE interpretation at 24 Months

A total of 4 studies were analyzed, which showed odds ratio of MACE was -0.30 (95% CI: -0.67; 0.07). The Highest odds of MACE were in 0.42 (95% CI: -0.61; 1.44). The lowest odds ratio was -1.42 (95% CI: -2.56; 1.44). The Heterogeneity was 1.96 and I-Squared was 48.98. The Forest Plot in Figure 9.

### 3.10 Change of Infarct size at 6 months

The total of 9 studies were analyzed, which showed MD of 1.80 (95% CI: 0.58; 3.02), The Highest MD of infarct size, Chulikana et.al. 2014, 6.60 (95% CI: -7.12, 20.32) and the lowest was wohrle et. Al. 2013 0.20 (95% CI: -9.21; 9.61) and Herbots et. Al. 2009, 0.20 (95% CI: -1.17; 1.57). The heterogeneity was 1.96 and I-Square was 91.09%.

### 3.11 Change of Infarct size at 12 months

The total of 10 studies were analyzed, which showed MD of 0.70 (95% CI: 0.48; 0.91), The Highest MD of infarct size, Yao et. Al. 2009, 3.12 (95% CI: -0.80, 7.04) and the lowest was wohrle et. Al. 2013 0.25 (95% CI: -0.14; 0.64). The heterogeneity was 0.06 and I-Square was 95.92%.

## 4. Discussion

Acute myocardial infarction (AMI) is a leading cause of morbidity and mortality worldwide, and its progression to heart failure (HF) remains a significant clinical challenge. Despite advances in early reperfusion strategies and pharmacological interventions, patients with AMI often experience persistent cardiac dysfunction, leading to an increased incidence of heart failure and poor long-term outcomes. The need for novel therapeutic strategies to mitigate this progression is paramount. Stem cell therapy has emerged as a promising treatment for myocardial repair, with numerous studies demonstrating its potential to improve cardiac function post-AMI. However, the clinical translation of these promising results remains hindered by a range of factors, including the optimal timing and dosage of stem cell administration, as well as the variability in study outcomes.

The present systematic review aimed to assess the mid-to long-term efficacy of stem cell therapy in AMI patients, focusing on changes in left ventricular ejection fraction (LVEF), infarct size, and major adverse cardiovascular events (MACE) at various time intervals. Our findings confirm the potential of stem cell therapy in improving cardiac function, but also highlight the challenges that remain in establishing consistent and robust clinical outcomes.

### 4.1 Efficacy of Stem Cell Therapy in AMI

The results of the analysis demonstrated an overall improvement in LVEF following stem cell therapy, with significant differences observed at various time points, including 6, 12, and 24 months. The pooled mean difference in LVEF at 6 months was 1.83%, and at 12 months, it increased to 2.21%. These findings are consistent with prior studies that have shown the regenerative potential of stem cells, such as mesenchymal stem cells (MSCs), induced pluripotent stem cells (iPSCs), and cardiac progenitor cells (CPCs), in improving myocardial function. However, the heterogeneity of the studies included in the analysis suggests that results can be inconsistent, as reflected by the large variations in LVEF improvements across individual studies. For example, studies by Wang et al. (2014) and Lezo et al. (2007) showed substantial improvements in LVEF, while others like Wohrle et al. (2010) found negligible or even negative effects. These discrepancies underscore the importance of addressing critical variables, including stem cell type, administration protocol, and patient-specific factors, which can all influence the therapeutic efficacy of stem cell therapy.

The mechanism underlying the improvement in cardiac function likely involves stem cells’ ability to differentiate into cardiomyocytes, support angiogenesis, and secrete paracrine factors that promote tissue repair. However, the long-term benefits of stem cell therapy remain uncertain, as the clinical outcomes at 24 and 36 months showed only modest improvements in LVEF. The lack of sustained benefits over time raises questions regarding the long-term viability and functional integration of stem cells into the damaged myocardial tissue [89]. This necessitates further investigation into how stem cells interact with the host tissue over extended periods and the factors that may contribute to the decline in therapeutic efficacy over time.

### 4.2 Impact on Infarct Size and MACE

In terms of infarct size, the analysis revealed a reduction at 6 months, with a mean difference of 1.80%. These findings align with the regenerative effects of stem cells on myocardial tissue, where reduced infarct size suggests potential for tissue regeneration [90]. However, at 12 months, the reduction in infarct size was minimal, indicating that the early benefits of stem cell therapy may not be sustained in the long term [91]. This observation emphasizes the need for improved delivery methods, such as the use of biomaterials or controlled-release systems, to enhance the retention and integration of stem cells in the infarcted area.

The analysis of major adverse cardiovascular events (MACE), which includes cardiovascular death, reinfarction, stroke, and hospitalization for heart failure, provided further insight into the clinical relevance of stem cell therapy. The pooled odds ratio for MACE at 6 months was -0.89, indicating a favorable trend for stem cell therapy in reducing adverse events. [92] However, the lack of significant improvements in MACE outcomes at 12, 24, and 36 months suggests that while stem cell therapy may provide short-term protection, its long-term impact on cardiovascular morbidity remains unclear. The role of stem cells in preventing recurrent infarctions or other adverse events needs further exploration, particularly in light of the heterogeneity observed in the MACE outcomes [93].

### 4.3 Challenges and Future Directions

Despite the promising results observed in the short to mid-term, several challenges persist in translating stem cell therapy into routine clinical practice. One of the major challenges is the lack of consensus on the optimal timing and dosage of stem cell administration. Early studies suggest that early administration may lead to better functional recovery, but the risks of arrhythmias and immune rejection remain significant. Conversely, late administration may limit the regenerative potential of stem cells due to extensive myocardial damage. Future studies must focus on determining the ideal window for stem cell intervention to maximize therapeutic benefits while minimizing risks.

Additionally, the type of stem cells used and the source of these cells play a crucial role in determining therapeutic outcomes. Allogeneic stem cells offer the advantage of immediate availability, but they carry the risk of immune rejection, whereas autologous stem cells avoid this issue but are time-consuming and costly to prepare. The heterogeneity in stem cell sources across the studies included in this review may partly explain the variability in outcomes.

The use of major adverse cardiovascular events (MACE) as a primary endpoint is a significant strength of this study, as it provides a more comprehensive assessment of clinical outcomes compared to traditional measures of cardiac function. Future trials should continue to prioritize MACE as a key endpoint to evaluate the true efficacy of stem cell therapy in preventing long-term cardiovascular complications. Furthermore, the incorporation of advanced imaging techniques, such as cardiac MRI and positron emission tomography (PET), may improve the accuracy of outcome measures and help refine the understanding of the mechanisms underlying the effects of stem cell therapy [94].

## 5. Conclusion

Stem cell therapy holds considerable promise for improving cardiac function and reducing the progression to heart failure following acute myocardial infarction. However, significant challenges remain in optimizing treatment protocols, including the timing, dosage, and stem cell source. While short-term improvements in LVEF and infarct size are encouraging, the long-term efficacy of stem cell therapy remains inconclusive. Future research should focus on large-scale, well-designed clinical trials that assess both the immediate and extended benefits of stem cell therapy, with particular attention to the use of MACE as a primary endpoint. By addressing these challenges, stem cell therapy may eventually become a cornerstone of AMI treatment, offering the potential for myocardial regeneration and improved patient outcomes.

## Authors’ contributions

All authors contributed equally to the manuscript and read and approved the final version of the manuscript.

## Data Availability

Supplementary file

---

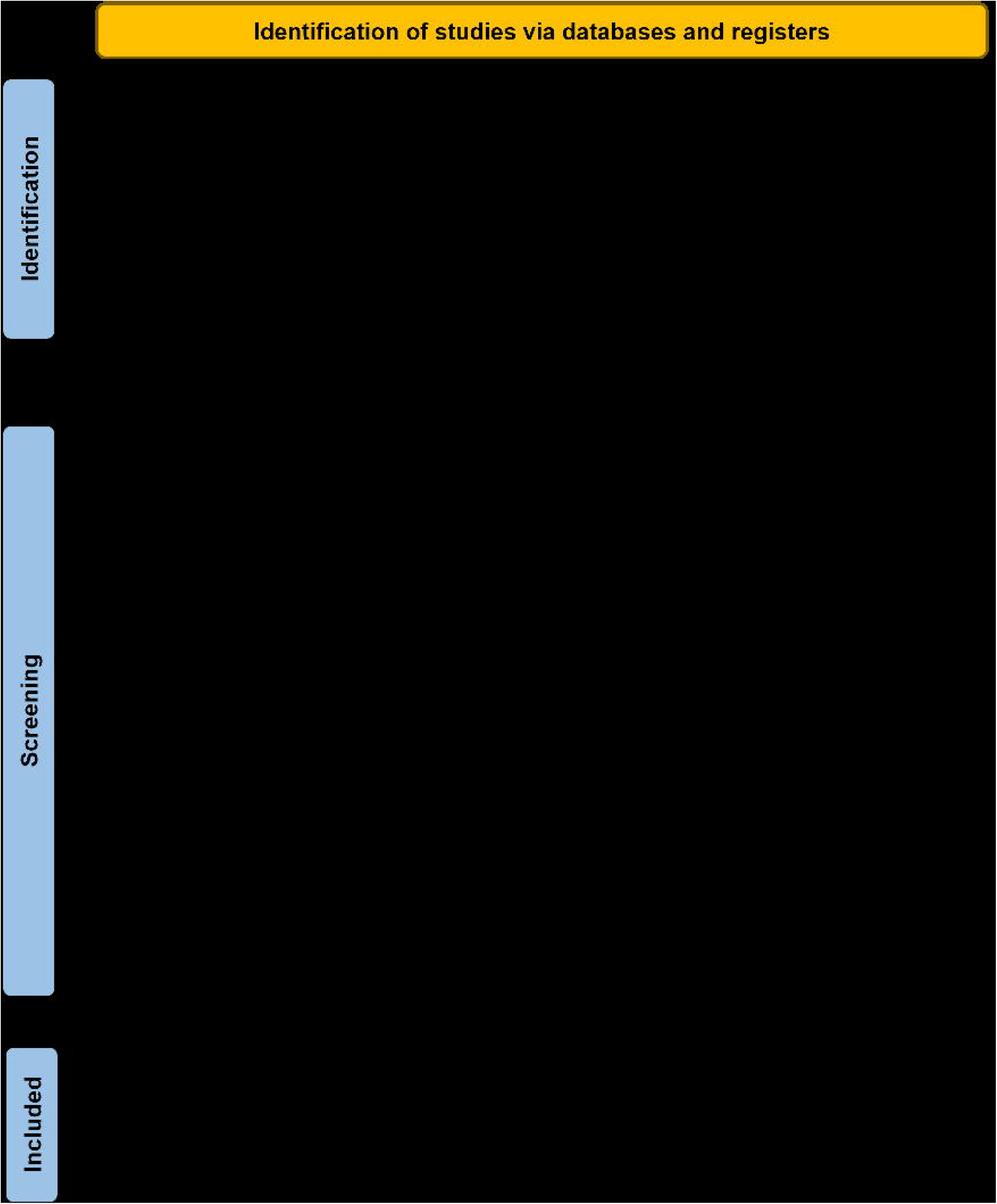

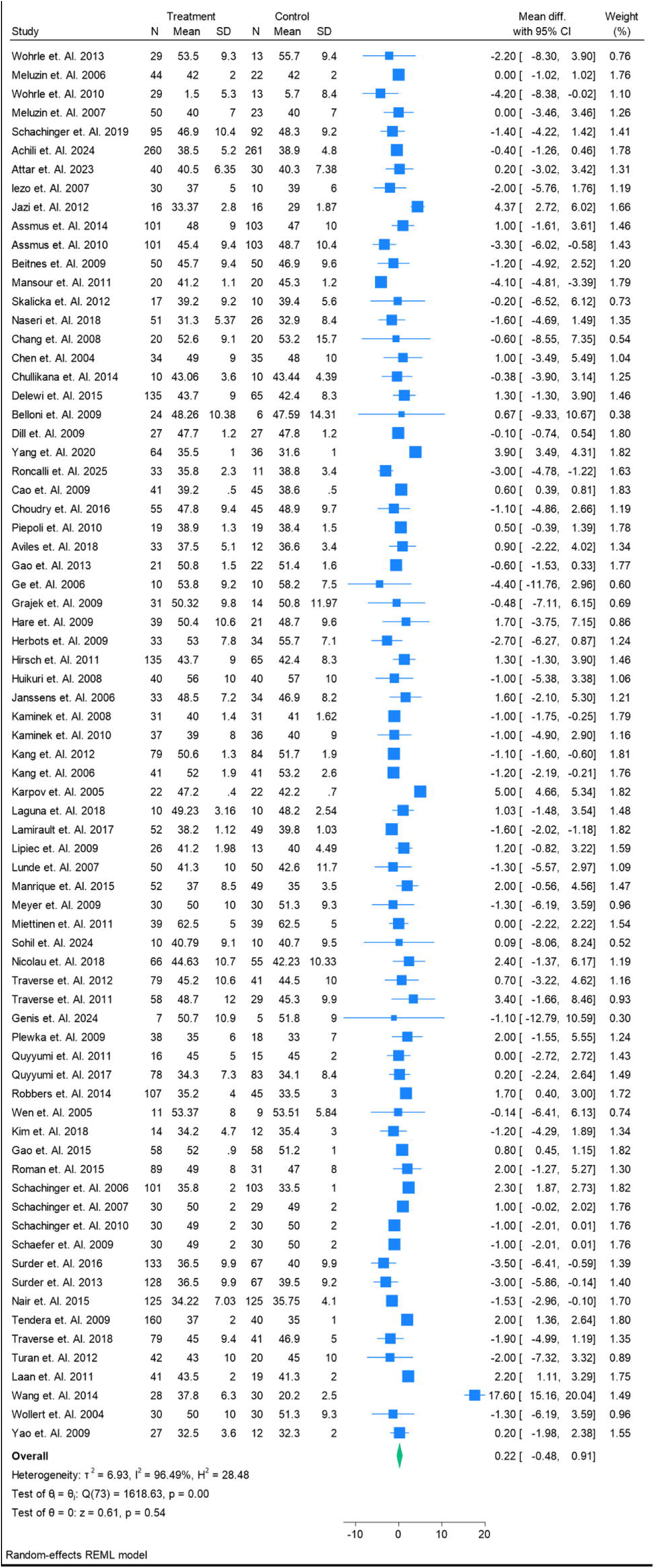

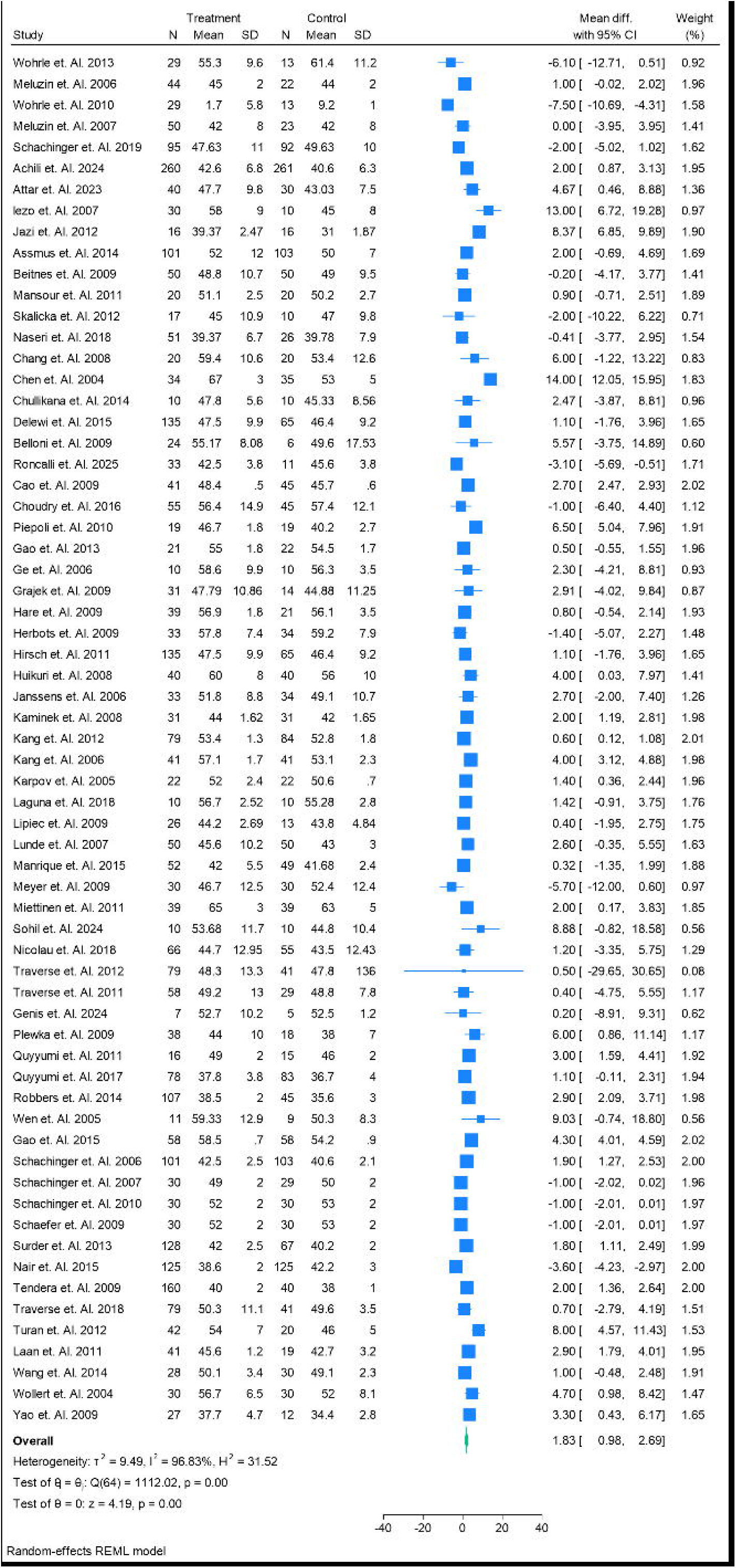

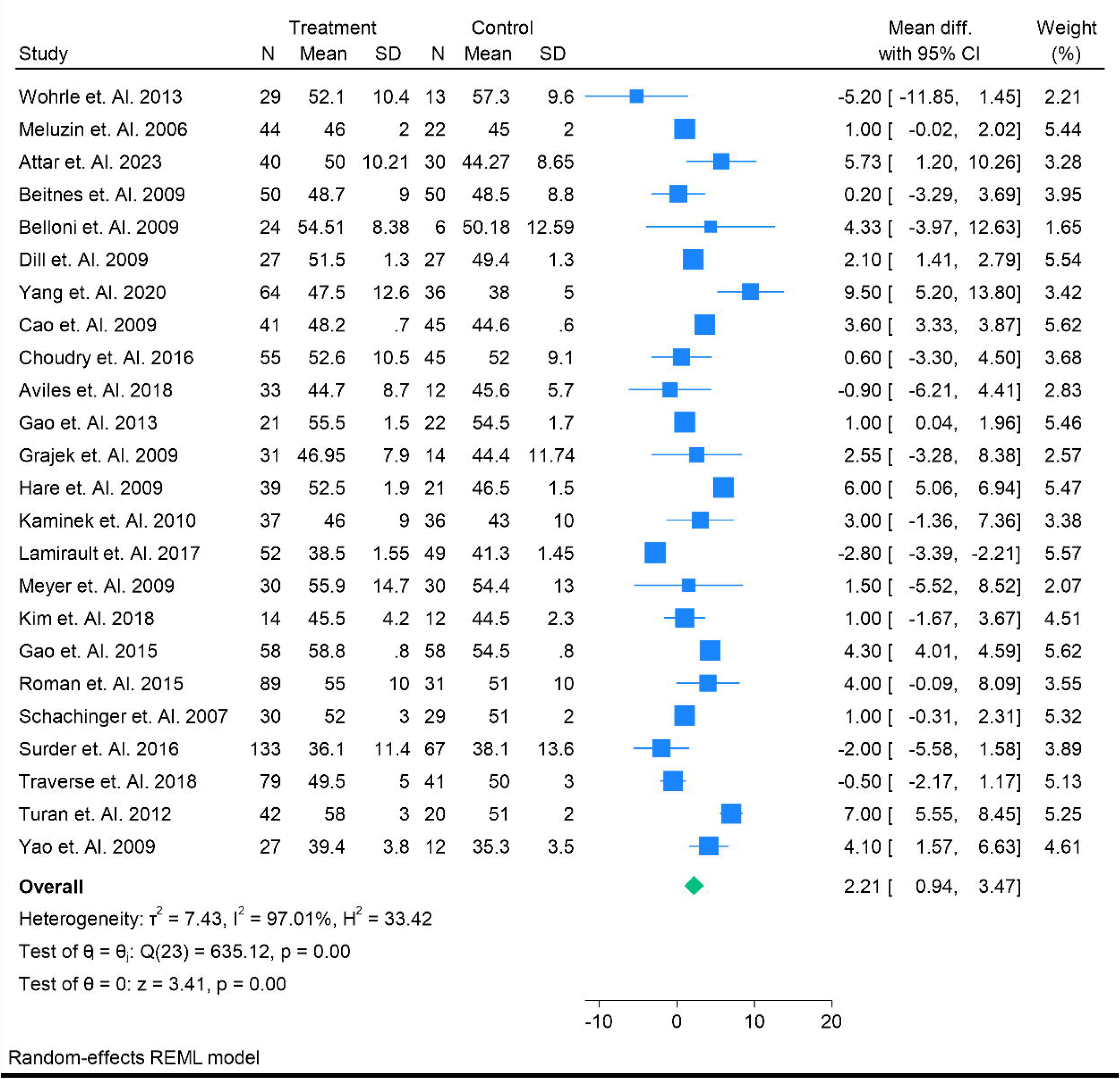

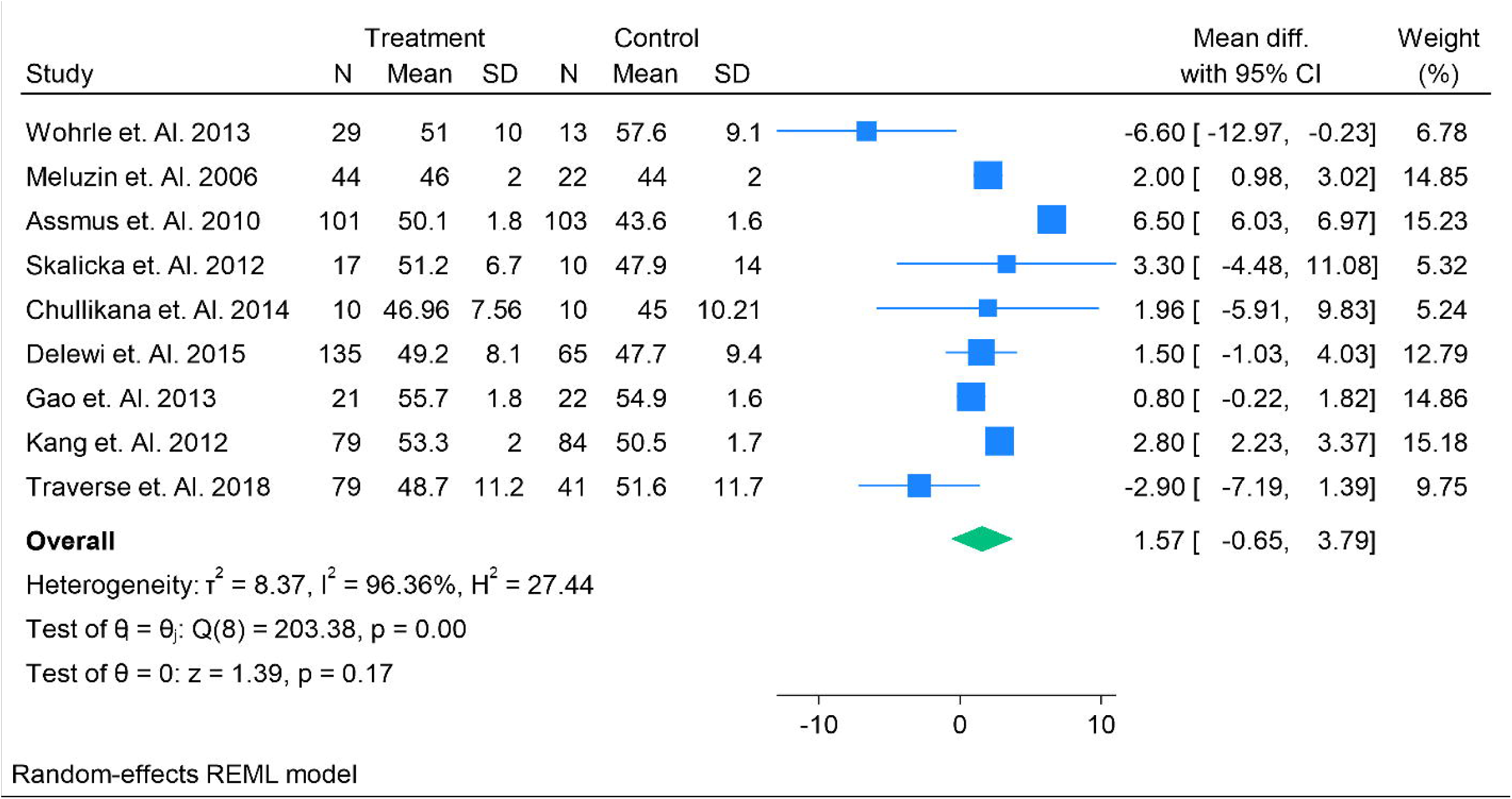

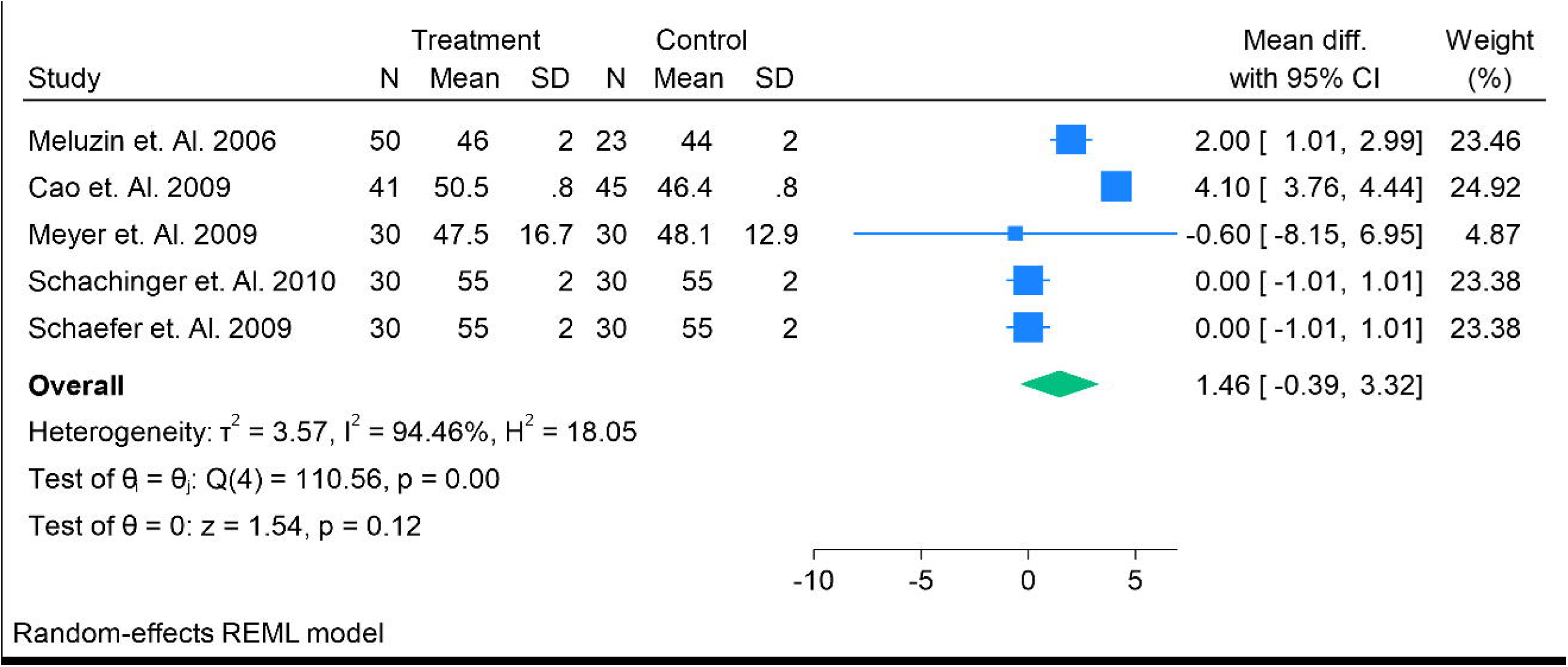

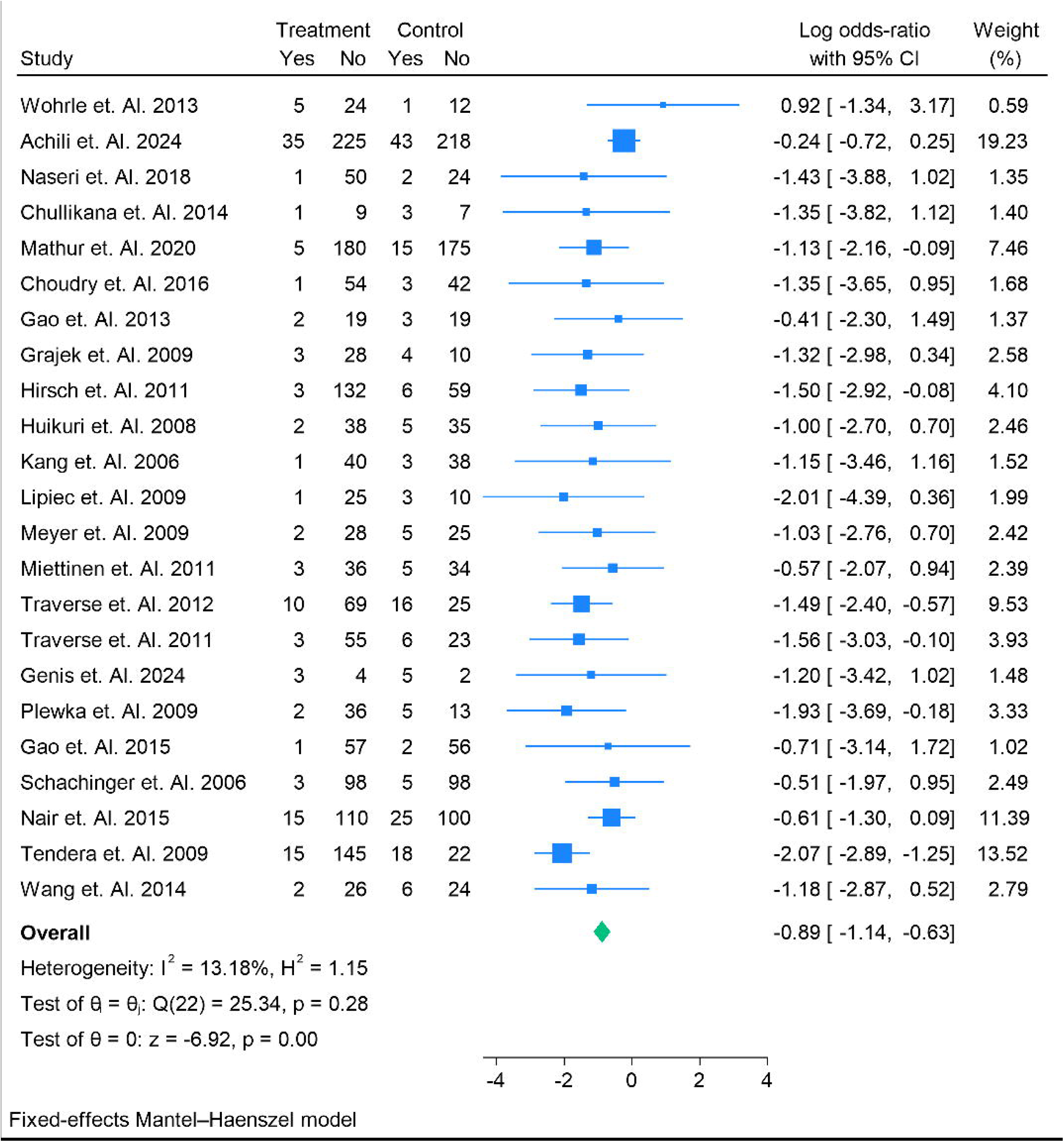

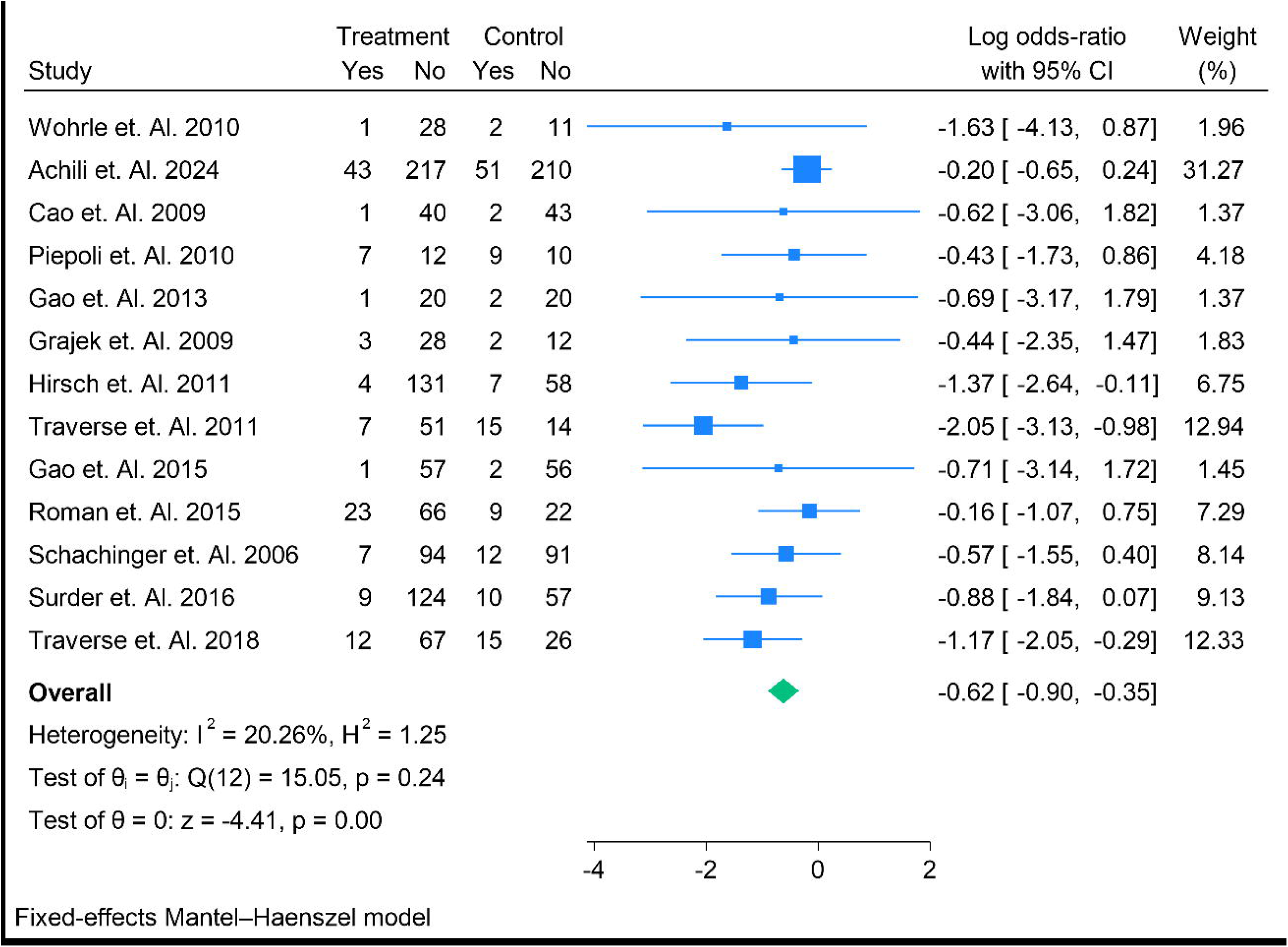

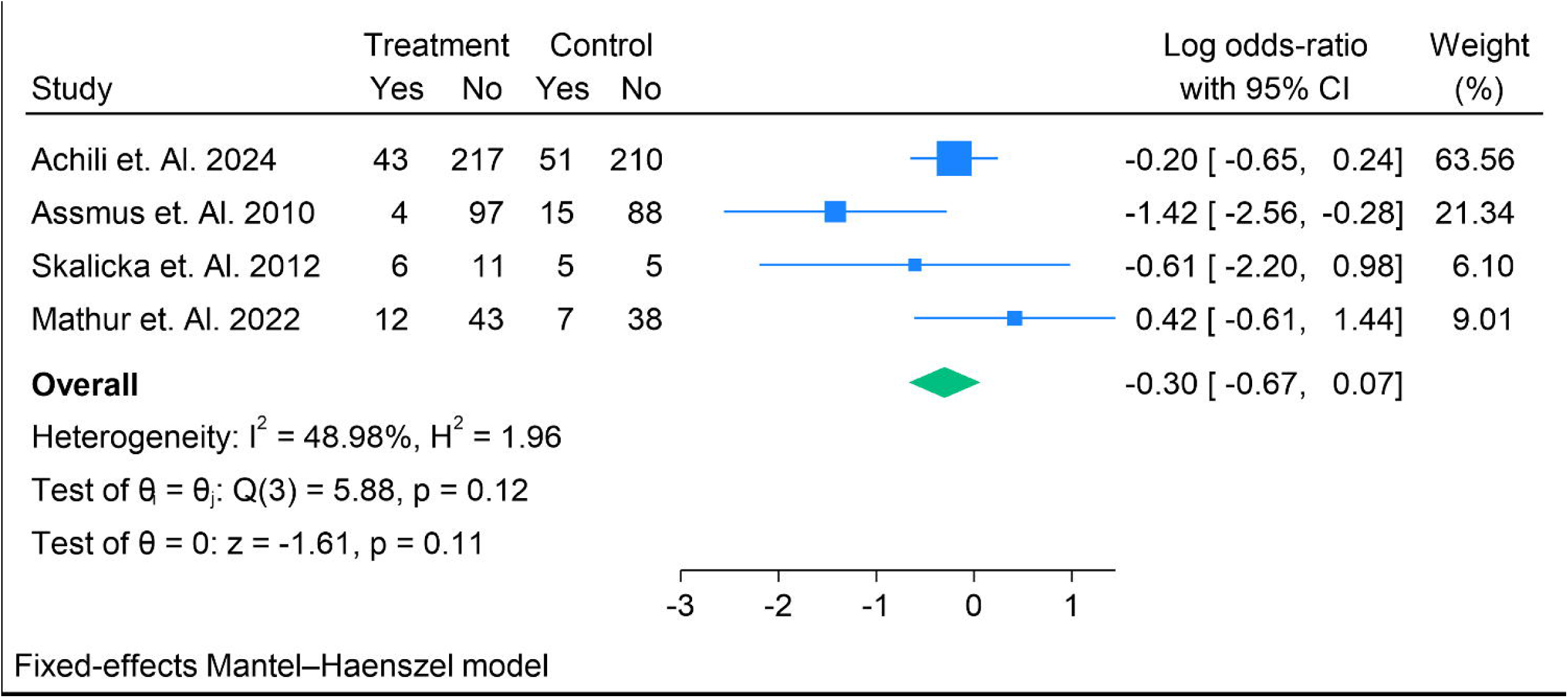

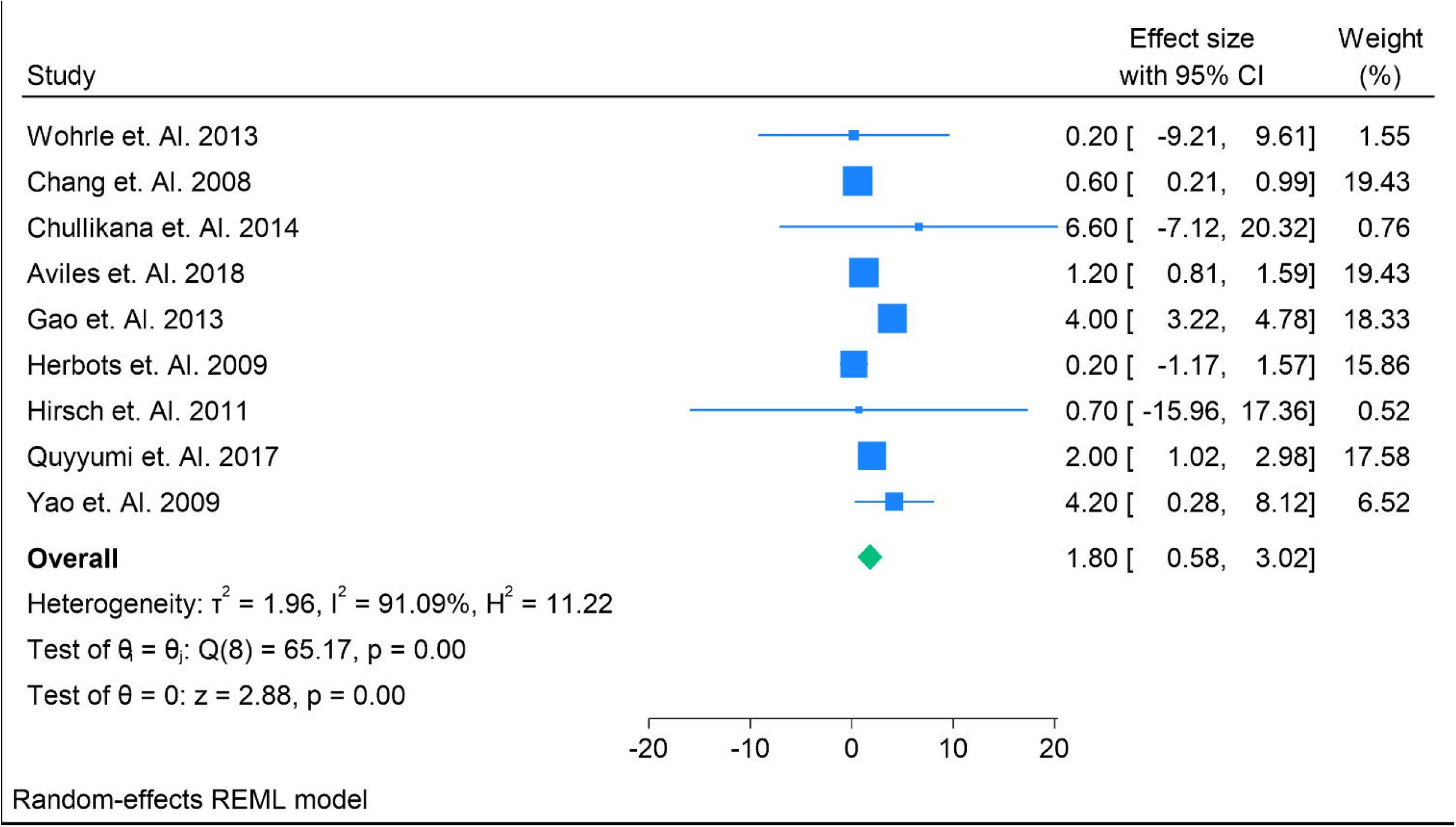

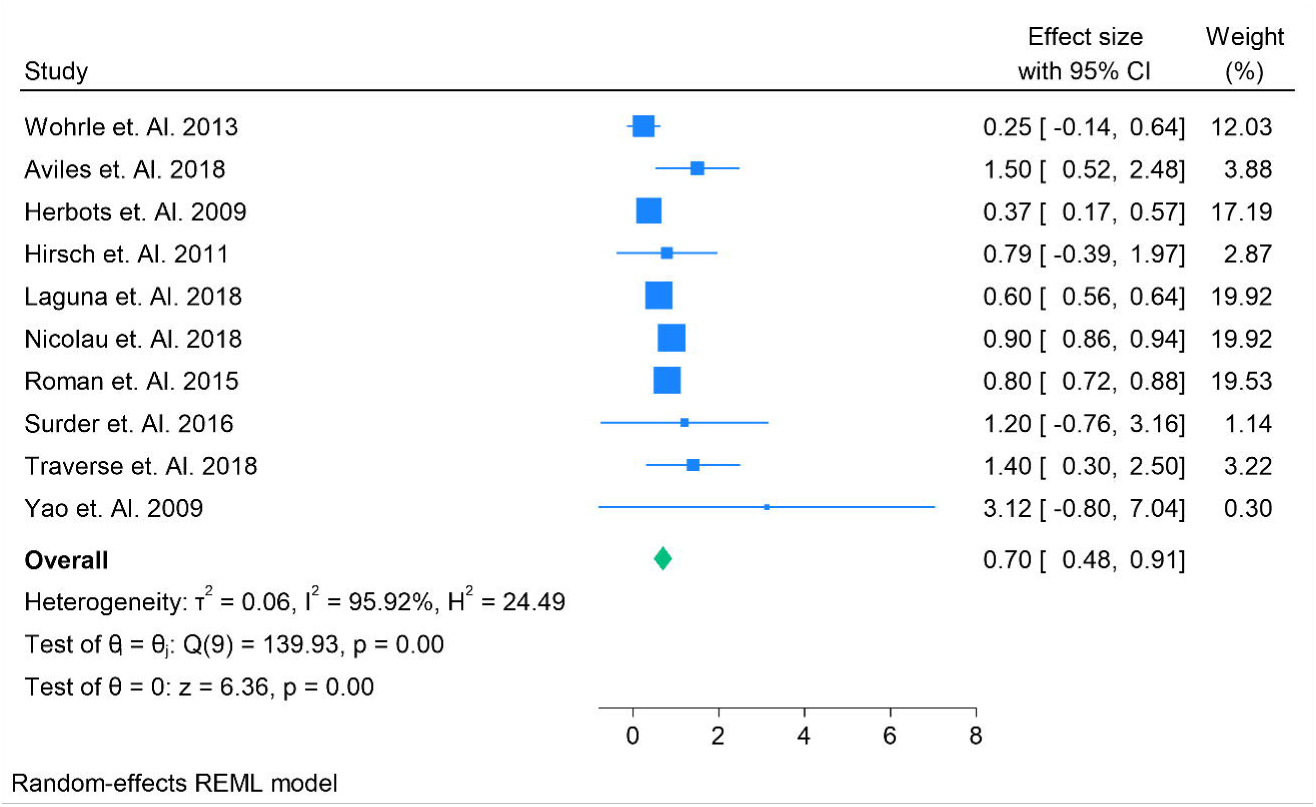

