## supplementary file for "Systematic Review and Meta-Analysis of Stem Cell Therapy in Myocardial Infarction: Effects on Left Ventricular Ejection Fraction and Major Adverse Cardiovascular Events"

Supplementary Files

Search String

((("myocardial infarction"[Mesh] OR "ST elevation myocardial infarction" OR "non-ST elevated myocardial infarction" OR "angina pectoris"[Mesh] OR "myocardial ischemia"[Mesh] OR "coronary artery disease"[Mesh] OR "coronary occlusion"[Mesh] OR "coronary stenosis"[Mesh] OR "acute coronary syndrome"[Mesh] OR STEMI OR NSTEMI) AND ("stem cells"[Mesh] OR "bone marrow cells"[Mesh] OR "mesenchymal stem cells"[Mesh] OR "mononuclear cells"[Mesh] OR "mesenchymal stromal cells" OR "pluripotent stromal cells" OR "embryonic stromal cells" OR "cardiac progenitor cells")) AND ("ejection fraction" OR "LVEF" OR "major adverse cardiac events" OR "MACE" OR "infarct size"))

Table S1. Demographics and Summary Table of All Studies

| **Ref No** | **Authors Year** | **Counrty of Trial** | **Stem Cell Origin** | **Type of Cell** | **Total Sample** | **Male** | **Female** | **Treatment Name** | **Rx No** | **Cx Name** | **Cx No** | **Mean Age** | **Route of Delivery** | **Cell Dosage** | **Duration of Culture** | **Injection Time** | **Follow Up** | **Grade** | **Main Finding** |
| --- | --- | --- | --- | --- | --- | --- | --- | --- | --- | --- | --- | --- | --- | --- | --- | --- | --- | --- | --- |
| 12 | Ramaseshan et. Al. 2024 | UK | Autologous Bone Marrow | BMMNCs | 110 | 55 | 55 | BMMNCs | 55 | Control | 55 |  | IC | 10 | 1 | 1 | 12 | High | The main finding of the REGENERATE-COBRA trial is that intracoronary infusion of autologous bone marrow-derived mononuclear cells in patients with refractory angina shows potential as a safe and effective treatment for improving cardiac function and symptoms. |
| 13 | Wohrle et. Al. 2013 | Germany | Bone Marrow | BMMNCs | 42 | 21 | 21 | BMMNCs | 29 | Control | 13 | 62 | IC | 32.4 | 1 | 1 | 36 | High | The main finding of the SCAMI trial is that higher doses of bone marrow-derived mononuclear cells (BMC) and the absence of microvascular obstruction (MO) led to a significant improvement in left ventricular ejection fraction (LVEF) in patients with acute myocardial infarction. |
| 14 | Meluzin et. Al. 2006 | Czech Republic | Bone Marrow | BMMNCs | 66 | 61 | 5 | BMMNCs | 44 | Control | 22 | 55 | IC | 0.9 | 1 | 9 | 3 | High | The main finding of the study is that autologous mononuclear bone marrow cell transplantation improves regional myocardial function in a dose-dependent manner in patients with acute myocardial infarction. |
| 15 | Wohrle et. Al. 2010 | Germany | Bone Marrow | BMMNCs | 42 | 36 | 6 | BMMNCs | 29 | Control | 13 | 36 | IC | 38.1 | 1 | 7 | 6 | High | The main finding of the study is that intracoronary bone marrow cell therapy did not show a significant improvement in left ventricular ejection fraction or infarct size compared to placebo in patients with acute myocardial infarction. |
| 16 | Meluzin et. Al. 2007 | Czech Republic | Bone Marrow | BMMNCs | 73 | 55 | 5 | BMMNCs | 50 | Control | 23 | 55 | IC | 1 | 1 | 1 | 12 | High | The main finding of the study is that autologous mononuclear bone marrow cell transplantation improves global left ventricular function in patients with acute myocardial infarction, with sustained benefits at 12 months for the higher cell dose group. |
| 17 | Schachinger et. Al. 2019 | Germany | Bone Marrow | BMMNCs | 204 | 155 | 49 | BMMNCs | 95 | Control | 92 | 56 | IC | 1 | 1 | 6 | 4 | High | The main finding of the study is that intracoronary infusion of bone marrow-derived mononuclear cells significantly improves left ventricular remodelling and contractility in patients after acute myocardial infarction. |
| 18 | Achili et. Al. 2024 | Italy | Bone Marrow | Granulocytes | 532 | 439 | 93 | G-CSF | 260 | Control | 261 | 60.9 | SC | 5 | 1 | 1 | 24 | High | The main finding of the STEM-AMI OUTCOME trial is that early administration of granulocyte colony-stimulating factor (G-CSF) did not significantly reduce the primary composite outcome of mortality, reinfarction, or heart failure hospitalization in patients with STEMI and left ventricular dysfunction, despite showing some trends in certain subgroups. |
| 19 | Attar et. Al. 2023 | Iran | Whartons Jelly | MSCs | 70 | 58 | 12 | MSCs | 40 | Control | 30 | 55.4 | IC | 1 | 1 | 10 | 6 | High | The main finding of the study is that intracoronary transplantation of Wharton’s jelly-derived mesenchymal stromal cells (WJ-MSCs) significantly improves left ventricular ejection fraction (LVEF) in patients after acute myocardial infarction, with a booster dose enhancing the effect. |
| 20 | lezo et. Al. 2007 | Spain | Bone Marrow | BMMNCs | 40 | 24 | 16 | BMMNCs | 30 | Control | 10 | 52 | IC | 90 | 1 | 1 | 7 | High | The main finding of the study is that intracoronary bone marrow cell therapy did not show a significant improvement in left ventricular ejection fraction or infarct size compared to placebo in patients with acute myocardial infarction. |
| 21 | Jazi et. Al. 2012 | Iran | Bone Marrow | BMMNCs | 32 | 25 | 7 | BMMNCs | 16 | Control | 16 | 45.2 | IC | 2.4 | 1 | 1 | 6 | High | Intracoronary infusion of autologous bone marrow progenitor cells in acute MI patients significantly improved LVEF over 6 months compared to control. |
| 22 | Assmus et. Al. 2014 | Germany | Bone Marrow | BMMNCs | 204 | 102 | 102 | BMMNCs | 101 | Control | 103 | 55 | IC | 1 | 1 | 7 | 61 | High | Intracoronary infusion of bone marrow-derived mononuclear cells after acute myocardial infarction improved long-term clinical outcomes, with better event-free survival linked to higher migratory capacity of the administered cells. |
| 23 | Assmus et. Al. 2010 | Germany | Bone Marrow | BMMNCs | 167 | 37 | 102 | BMMNCs | 101 | Control | 103 | 56 | IC | 1 | 1 | 7 | 24 | High | Intracoronary infusion of bone marrow–derived progenitor cells significantly reduced major adverse cardiovascular events and improved left ventricular function over 2 years in patients with acute myocardial infarction. |
| 24 | Beitnes et. Al. 2009 | Norway | Bone Marrow | BMMNCs | 100 | 84 | 16 | BMMNCs | 50 | Control | 50 | 58.1 | IC | 68.6 | 1 | 8 | 36 | High | Intracoronary injection of autologous mononuclear bone marrow cells after acute myocardial infarction was safe but did not improve left ventricular function or clinical outcomes over 3 years. |
| 25 | Mansour et. Al. 2011 | Canada | Bone Marrow | CD133+ | 40 | 36 | 4 | CD133+ | 20 | Control | 20 | 52.2 | IC | 1 | 1 | 6.4 | 12 | High | Intracoronary injection of selected CD133+ bone marrow stem cells in post-MI patients with LV dysfunction was safe and led to significant improvement in left ventricular ejection fraction at 12 months. |
| 26 | Skalicka et. Al. 2012 | Czech Republic | Bone Marrow | BMMNCs | 27 | 22 | 5 | BMMNCs | 17 | Control | 10 | 61 | IC | 0.13 | 1 | 9 | 24 | High | Intracoronary injection of autologous bone marrow-derived mononuclear cells resulted in a significant improvement in left ventricular ejection fraction at 24 months compared to standard therapy in patients with large anterior myocardial infarction. |
| 27 | Naseri et. Al. 2018 | Iran | Bone Marrow | BMMNCs | 77 | 69 | 8 | BMMNCs | 51 | Control | 26 | 53.3 | IC | 56.4 | 1 | 1 | 18 | High | Intramyocardial injection of autologous CD133⁺ cells significantly improved LVEF and reduced non-viable myocardial segments compared to placebo and MNCs in patients with recent MI undergoing CABG. |
| 28 | Chang et. Al. 2008 | South Korea | G-CSF | CD34+ | 40 | 33 | 7 | CD34+ | 20 | Control | 20 | 57 | IC | 0.7 | 1 | 3 | 6 | High | Stem cell therapy using G-CSF–mobilised peripheral blood stem cells significantly improved left ventricular ejection fraction and restored ventricular synchrony in patients with acute myocardial infarction at 6 months. |
| 29 | Chen et. Al. 2004 | China | Bone Marrow | BMMNCs | 69 | 66 | 3 | BMMNCs | 34 | Control | 35 | 58 | IC | 800 | 10 | 8 | 6 | High | Intracoronary transplantation of autologous bone marrow mesenchymal stem cells significantly improved left ventricular function in patients with acute myocardial infarction at 6-month follow-up. |
| 30 | Chullikana et. Al. 2014 | India | Bone Marrow | BMMSCs | 20 | 18 | 2 | BMMSCs | 10 | Control | 10 | 47.5 | IV | 2 | 4 | 2 | 24 | High | Intravenous administration of allogeneic bone marrow-derived mesenchymal stromal cells (Stempeucel) in acute myocardial infarction patients was safe but did not significantly improve cardiac function compared to placebo over 24 months. |
| 31 | Delewi et. Al. 2015 | Netherlands | Bone Marrow | BMMNCs | 200 | 170 | 30 | BMMNCs | 135 | Control | 65 | 56 | IC | 1.5 | 1 | 8 | 60 | High | Intracoronary infusion of bone marrow mononuclear cells (BMMC) after myocardial infarction modestly reduced left ventricular remodeling without improving long-term clinical outcomes, while peripheral blood mononuclear cells (PBMC) were associated with worse outcomes. |
| 32 | Belloni et. Al. 2009 | Brazil | Bone Marrow | BMMNCs | 30 | 21 | 9 | BMMNCs | 24 | Control | 6 | 59 | IV | 10 | 1 | 1 | 6 | High | Autologous mononuclear bone marrow cell transplantation did not significantly improve left ventricular systolic function in patients with acute myocardial infarction after 6 months. |
| 33 | Dill et. Al. 2009 | Germany | Bone Marrow | BMMNCs | 54 | 49 | 5 | BMMNCs | 27 | Control | 27 | 57.9 | IC | 1 | 1 | 8 | 12 | High | Intracoronary administration of bone marrow-derived progenitor cells significantly improved left ventricular function and limited adverse remodeling in post-STEMI patients with reduced baseline ejection fraction at 12 months. |
| 34 | Yang et. Al. 2020 | China | Bone Marrow | BMMNCs | 100 | 68 | 32 | BMMNCs | 64 | Control | 36 | 52.9 | IC | 1 | 1 | 1 | 12 | High | Intensive atorvastatin significantly enhances the therapeutic efficacy of autologous bone marrow mononuclear cell transplantation in improving left ventricular ejection fraction in patients with anterior STEMI. |
| 35 | Mathur et. Al. 2020 | Multicenter | Bone Marrow | BMMNCs | 375 | 302 | 73 | BMMNCs | 185 | Control | 190 | 59 | IC | 5 | 2 | 8 | 24 | High | Intracoronary infusion of autologous bone marrow-derived mononuclear cells after acute myocardial infarction was safe but did not significantly reduce all-cause mortality or major adverse cardiac events due to low recruitment and event rates. |
| 36 | Mathur et. Al. 2022 | Multicenter | Bone Marrow | BMMNCs | 100 | 76 | 24 | BMMNCs | 55 | Control | 45 | 56.6 | IC | 1 | 1 | 1 | 60 | High | Autologous intracoronary bone marrow cell therapy after acute myocardial infarction did not improve clinical outcomes at 5 years compared to placebo. |
| 37 | Roncalli et. Al. 2025 | France | Bone Marrow | CD34+ | 44 | 22 | 22 | CD34+ | 33 | Control | 11 | 52 | IC | 1 | 1 | 9 | 6 | High | The EXCELLENT trial is designed to evaluate the safety and feasibility of transendocardial injection of expanded autologous CD34+ cells (ProtheraCytes®) in patients with large myocardial infarction and reduced LVEF, with results pending. |
| 38 | Cao et. Al. 2009 | China | Bone Marrow | BMMNCs | 86 | 81 | 5 | BMMNCs | 41 | Control | 45 | 50.7 | IC | 12.5 | 1 | 7 | 48 | High | Intracoronary transplantation of autologous bone marrow mononuclear cells in STEMI patients significantly improved long-term left ventricular function over 4 years without major adverse effects. |
| 39 | Choudry et. Al. 2016 | UK | Bone Marrow | BMMNCs | 100 | 87 | 13 | BMMNCs | 55 | Control | 45 | 56.5 | IC | 5.9 | 1 | 1 | 12 | High | Early intracoronary infusion of autologous bone marrow cells after AMI resulted in a small, non-significant improvement in LVEF at 1 year but showed benefits in infarct size reduction and myocardial salvage. |
| 40 | Piepoli et. Al. 2010 | Italy | Bone Marrow | BMMNCs | 38 | 26 | 12 | BMMNCs | 19 | Control | 19 | 63.1 | IC | 41.8 | 1 | 7 | 12 | High | Intracoronary infusion of autologous bone marrow cells significantly improved left ventricular function, autonomic control, and exercise capacity in post-myocardial infarction patients over 12 months. |
| 41 | Aviles et. Al. 2018 | Spain | CSC | AlloCSC-01 | 49 | 45 | 4 | AlloCSC-01 | 33 | Control | 12 | 55 | IC | 1.2 | 1 | 7 | 12 | High | Intracoronary infusion of allogeneic cardiac stem cells (AlloCSC-01) in STEMI patients was safe but showed no significant benefit in reducing infarct size or improving left ventricular function at 12 months. |
| 42 | Gao et. Al. 2013 | China | Bone Marrow | BMMSCs | 43 | 40 | 3 | BMMSCs | 21 | Control | 22 | 55 | IC | 14.6 | 1 | 7 | 24 | High | Intracoronary injection of autologous bone marrow mesenchymal stem cells after acute myocardial infarction improved myocardial viability but did not significantly enhance cardiac function or reduce infarct size compared to standard therapy. |
| 43 | Ge et. Al. 2006 | China | Bone Marrow | BMMNCs | 20 | 18 | 2 | BMMNCs | 10 | Control | 10 | 58 | IC | 4 | 1 | 7 | 6 | High | Emergent intracoronary transplantation of autologous bone marrow mononuclear cells after acute myocardial infarction significantly improved left ventricular function and myocardial perfusion at 6 months without adverse events. |
| 44 | Grajek et. Al. 2009 | Poland | Bone Marrow | BMMNCs | 45 | 39 | 6 | BMMNCs | 31 | Control | 14 | 54 | IC | 41 | 1 | 7 | 12 | High | Intracoronary infusion of bone marrow stem cells after anterior wall myocardial infarction did not significantly improve ejection fraction but showed a modest enhancement in myocardial perfusion. |
| 45 | Hare et. Al. 2009 | USA | Bone Marrow | hMSCs | 53 | 43 | 10 | hMSCs | 39 | Control | 21 | 59 | IV | 0.5 | 1 | 10 | 24 | High | Intravenous infusion of allogeneic bone marrow-derived mesenchymal stem cells (Prochymal) after acute myocardial infarction was safe and showed potential efficacy by improving left ventricular function and reducing arrhythmias. |
| 46 | Herbots et. Al. 2009 | Belgium | Bone Marrow | BPMCs | 67 | 55 | 12 | BPMCs | 33 | Control | 34 | 53 | IC | 0.6 | 1 | 5 | 4 | High | Intracoronary infusion of autologous bone marrow progenitor cells significantly improved regional myocardial function in infarcted segments at 4 months, without affecting global LVEF. |
| 47 | Hirsch et. Al. 2011 | Netherlands | Bone Marrow | BMMNCs | 200 | 114 | 86 | BMMNCs | 135 | Control | 65 | 56 | IC | 29.6 | 1 | 7 | 4 | High | Intracoronary infusion of mononuclear cells from bone marrow or peripheral blood after acute myocardial infarction did not improve left ventricular function or reduce infarct size compared to standard therapy. |
| 48 | Huikuri et. Al. 2008 | Finland | Bone Marrow | BMMNCs | 80 | 70 | 10 | BMMNCs | 40 | Control | 40 | 60 | IC | 40.2 | 1 | 3 | 6 | High | Intracoronary injection of autologous bone marrow mononuclear cells significantly improved left ventricular ejection fraction at 6 months without increasing arrhythmia risk or restenosis in thrombolysis-treated STEMI patients. |
| 49 | Janssens et. Al. 2006 | Belgium | Bone Marrow | BMMNCs | 67 | 55 | 12 | BMMNCs | 33 | Control | 34 | 45 | IC | 30 | 1 | 2 | 16 | High | Intracoronary infusion of autologous bone marrow-derived stem cells after STEMI did not significantly improve global left ventricular function but reduced infarct size and improved regional systolic function at 4 months. |
| 50 | Kaminek et. Al. 2008 | Czech Republic | Bone Marrow | BMMNCs | 62 | 54 | 8 | BMMNCs | 31 | Control | 31 | 55 | IC | 10 | 1 | 9 | 3 | High | Intracoronary transplantation of autologous bone marrow mononuclear cells modestly improved left ventricular function at 3 months post-MI, particularly in patients with higher baseline myocardial perfusion. |
| 51 | Kaminek et. Al. 2010 | Czech Republic | Bone Marrow | BMMNCs | 73 | 64 | 9 | BMMNCs | 37 | Control | 36 | 54 | IC | 1 | 1 | 1 | 12 | High | Intracoronary transplantation of autologous bone marrow mononuclear cells significantly improved left ventricular function at 12 months in patients with moderate residual myocardial viability, but not in those with severely scarred myocardium. |
| 52 | Kang et. Al. 2012 | South Korea | Bone Marrow | BMMNCs | 163 | 135 | 28 | BMMNCs | 79 | Control | 84 | 57.5 | IC | 1.1 | 1 | 5 | 24 | High | Intracoronary infusion of G-CSF–mobilized peripheral blood stem cells improved left ventricular function and significantly reduced major adverse cardiac events over 5 years in myocardial infarction patients. |
| 53 | Kang et. Al. 2006 | South Korea | Peripheral Blood | G-CSF | 82 | 68 | 14 | G-CSF | 41 | Control | 41 | 60 | IC | 0.7 | 1 | 2 | 6 | High | Intracoronary infusion of G-CSF–mobilized peripheral blood stem cells significantly improved left ventricular function and reduced infarct size in patients with acute myocardial infarction but showed no significant benefit in those with old myocardial infarction. |
| 54 | Karpov et. Al. 2005 | Russia | Bone Marrow | BMMNCs | 44 | 36 | 4 | BMMNCs | 22 | Control | 22 | 55.2 | IC | 0.2 | 1 | 21 | 6 | High | Intracoronary injection of autologous bone marrow mononuclear cells after acute myocardial infarction was safe, reduced inflammatory markers, but did not significantly improve left ventricular function over 6 months. |
| 55 | Laguna et. Al. 2018 | Spain | Bone Marrow | BMMNCs | 20 | 19 | 1 | BMMNCs | 10 | Control | 10 | 62.5 | IC | 1 | 1 | 15 | 9 | High | Intramyocardial autologous BMMNC grafting during CABG in subacute AMI showed no significant improvement in LVEF or ventricular remodeling at 9 months compared to CABG alone. |
| 56 | Lamirault et. Al. 2017 | France | Bone Marrow | BMMNCs | 101 | 51 | 50 | BMMNCs | 52 | Control | 49 | 62 | IC | 1 | 1 | 7 | 12 | High | Intracoronary injection of autologous bone marrow cells after acute myocardial infarction significantly improved quality of life at 3 and 12 months without enhancing cardiac function. |
| 57 | Lipiec et. Al. 2009 | Poland | Bone Marrow | BMMNCs | 36 | 25 | 11 | BMMNCs | 26 | Control | 13 | 57 | IC | 2.2 | 1 | 11 | 6 | High | Intracoronary injection of autologous mononuclear bone marrow cells in patients with acute myocardial infarction significantly improved myocardial perfusion at 6 months, with modest benefit to infarct-area systolic function but no clear enhancement of global left ventricular function. |
| 58 | Lunde et. Al. 2007 | Norway | Bone Marrow | BMMNCs | 100 | 84 | 16 | BMMNCs | 50 | Control | 50 | 58 | IC | 6.8 | 1 | 6 | 6 | High | Intracoronary injection of autologous mononuclear bone marrow cells after acute myocardial infarction significantly improved exercise time and heart rate response, but not peak oxygen consumption or quality of life, at 6-month follow-up. |
| 59 | Manrique et. Al. 2015 | France | Bone Marrow | BMMNCs | 101 | 51 | 50 | BMMNCs | 52 | Control | 49 | 51 | IC | 1.5 | 1 | 4 | 12 | High | LV remodelling 1 year after reperfused MI is associated with progressive dyssynchrony and driven by baseline infarct size and ejection fraction, with no significant impact from bone marrow cell therapy. |
| 60 | Meyer et. Al. 2009 | Germany | Bone Marrow | BMMNCs | 60 | 30 | 30 | BMMNCs | 30 | Control | 30 | 51 | IC | 1.5 | 1 | 5 | 12 | High | A single intracoronary infusion of autologous bone marrow cells (BMCs) did not provide sustained improvement in left ventricular ejection fraction (LVEF) after 5 years in STEMI patients, though a subgroup with more transmural infarcts showed potential long-term benefit. |
| 61 | Miettinen et. Al. 2011 | Finland | Bone Marrow | BMMNCs | 80 | 40 | 40 | BMMNCs | 39 | Control | 39 | 60 | IC | 40 | 1 | 1 | 6 | High | Intracoronary injection of autologous bone marrow-derived stem cells (BMC) did not significantly affect natriuretic peptide levels or inflammatory markers in STEMI patients compared to placebo. |
| 62 | Sohil et. Al. 2024 | India | Peripheral Blood | PBMNCs | 20 | 20 | 0 | PBMNCs | 10 | Control | 10 | 58 | IC | 1 | 1 | 7 | 6 | High | Intracoronary transplantation of autologous peripheral blood-derived mononuclear cells (PBMNCs) significantly improved left ventricular ejection fraction (LVEF) and wall motion score index (WMSI) in acute myocardial infarction patients compared to controls, with no adverse events reported. |
| 63 | Nicolau et. Al. 2018 | Brazil | Bone Marrow | BMMNCs | 121 | 93 | 28 | BMMNCs | 66 | Control | 55 | 59.02 | IC | 1 | 1 | 2 | 6 | High | Intracoronary infusion of bone marrow-derived mononuclear cells (BMMC) did not improve left ventricular ejection fraction (LVEF) or reduce infarct size in STEMI patients with reduced ejection fraction at 6-month follow-up. |
| 64 | Traverse et. Al. 2012 | USA | Bone Marrow | BMMNCs | 120 | 105 | 15 | BMMNCs | 79 | Control | 41 | 56.9 | IC | 15 | 1 | 7 | 6 | High | Intracoronary delivery of autologous bone marrow mononuclear cells (BMCs) at 3 or 7 days post-STEMI did not improve left ventricular function compared to placebo at 6 months. |
| 65 | Traverse et. Al. 2011 | USA | Bone Marrow | BMMNCs | 87 | 72 | 15 | BMMNCs | 58 | Control | 29 | 52.1 | IC | 1 | 1 | 7 | 24 | High | The LateTIME trial found no significant improvement in LV function with delayed (2–3 weeks) intracoronary BMC delivery post-MI. The study was well-designed with low risk of bias across all domains. |
| 66 | Genis et. Al. 2024 | Spain | Whartons Jelly | WJ-MSCs | 12 | 9 | 3 | BMMNCs | 7 | Control | 5 | 63.9 | IC | 0.7 | 1 | 7 | 12 | High | The PeriCord graft demonstrated safety and immunomodulatory properties in patients with myocardial infarction but showed no significant improvement in cardiac function or scar size at one-year follow-up. |
| 67 | Plewka et. Al. 2009 | Poland | Bone Marrow | BMMNCs | 60 | 42 | 18 | BMMNCs | 38 | Control | 18 | 56 | IC | 0.3 | 1 | 7 | 6 | High | Intracoronary injection of autologous bone marrow stem cells (BMSCs) significantly improved left ventricular systolic and diastolic function at 6 months in patients with acute myocardial infarction compared to controls. |
| 68 | Quyyumi et. Al. 2011 | USA | Bone Marrow | BMMNCs | 31 | 27 | 4 | BMMNCs | 16 | Control | 15 | 52 | IC | 5 | 1 | 5 | 12 | High | Infusion of ≥10 million autologous CD34+ cells into the infarct-related artery after STEMI significantly improved myocardial perfusion and showed a trend toward enhanced cardiac function at 6 months, with efficacy dependent on cell dose and mobility. |
| 69 | Quyyumi et. Al. 2017 | USA | Bone Marrow | BMMNCs | 161 | 132 | 29 | BMMNCs | 78 | Control | 83 | 56.4 | IC | 1 | 2 | 10 | 18 | High | Intracoronary infusion of autologous CD34+ cells in post-STEMI patients with left ventricular dysfunction was safe and showed potential efficacy, with higher cell doses associated with improved outcomes (e.g., LVEF, infarct size reduction, and reduced mortality). |
| 70 | Robbers et. Al. 2014 | Netherlands | Bone Marrow | BMMNCs | 152 | 131 | 21 | BMMNCs | 107 | Control | 45 | 56 | IC | 0.2 | 1 | 5 | 4 | High | Intracoronary infusion of bone marrow-derived or peripheral blood-derived mononuclear cells did not improve myocardial perfusion recovery in the infarct core or border zone after revascularization of ST-elevation myocardial infarction. |
| 71 | Wen et. Al. 2005 | China | Bone Marrow | BMMNCs | 20 | 19 | 1 | BMMNCs | 11 | Control | 9 | 58 | IC | 0.4 | 1 | 7 | 6 | High | Autologous bone marrow stem cell transplantation improved left ventricular function and reduced remodeling in acute myocardial infarction patients compared to controls. |
| 72 | Kim et. Al. 2018 | South Korea | Bone Marrow | BMMNCs | 26 | 26 | 0 | BMMNCs | 14 | Control | 12 | 57.8 | IC | 7.2 | 25 | 30 | 12 | High | Intracoronary autologous BM-MSC therapy significantly improved left ventricular ejection fraction (LVEF) at 4 and 12 months in patients with anterior STEMI, without increasing adverse events. |
| 73 | Gao et. Al. 2015 | China | Whartons Jelly | WJ-MSCs | 116 | 106 | 10 | WJ-MSCs | 58 | Control | 58 | 56.7 | IC | 0.6 | 1 | 7 | 18 | High | Intracoronary infusion of Wharton’s jelly-derived mesenchymal stem cells (WJMSCs) significantly improved myocardial viability, perfusion, and left ventricular function in patients with acute myocardial infarction, demonstrating safety and efficacy over 18 months. |
| 74 | Roman et. Al. 2015 | Spain | Bone Marrow | BMMNCs | 120 | 107 | 13 | BMMNCs | 89 | Control | 31 | 56 | IC | 8.3 | 3 | 5 | 12 | High | The study found no significant improvement in left ventricular ejection fraction (LVEF) or volumes with bone marrow-derived stem cell therapies (BMMC, G-CSF, or combined) compared to standard care in reperfused STEMI patients at 12-month follow-up. |
| 75 | Schachinger et. Al. 2006 | Germany | Bone Marrow | BMMNCs | 204 | 166 | 38 | BMMNCs | 101 | Control | 103 | 57 | IC | 1 | 2 | 7 | 12 | High | Intracoronary administration of bone-marrow-derived progenitor cells (BMCs) significantly reduced major adverse cardiovascular events (MACE) at 1 year in patients with acute myocardial infarction. |
| 76 | Schachinger et. Al. 2007 | Germany | Bone Marrow | BMMNCs | 60 | 41 | 18 | BMMNCs | 30 | Control | 29 | 59 | IC | 250 | 3 | 7 | 18 | High | Intracoronary autologous bone marrow cell transfer improved diastolic function parameters (E/A and Ea/Aa ratios) in patients after acute myocardial infarction. |
| 77 | Schachinger et. Al. 2010 | Germany | Bone Marrow | BMMNCs | 60 | 30 | 30 | BMMNCs | 30 | Control | 30 | 56 | IC | 1 | 4.8 | 5 | 60 | High | Intracoronary autologous bone marrow cell transfer improved diastolic function early (at 6 and 18 months) but did not sustain this benefit at 5-year follow-up in post-myocardial infarction patients. |
| 78 | Schaefer et. Al. 2009 | Germany | Bone Marrow | BMMNCs | 60 | 30 | 30 | BMMNCs | 30 | Control | 30 | 56 | IC | 2.5 | 4 | 5 | 60 | High | Intracoronary autologous bone marrow cell transfer improved diastolic function early (up to 18 months) but did not sustain benefits at 5-year follow-up in post-MI patients. |
| 79 | Surder et. Al. 2016 | Switzerland | Bone Marrow | BMMNCs | 200 | 100 | 100 | BMMNCs | 133 | Control | 67 | 62 | IC | 15 | 2 | 5 | 60 | High | Intracoronary infusion of bone marrow-derived mononuclear cells (BM-MNC) early (5–7 days) or late (3–4 weeks) after acute myocardial infarction did not improve left ventricular function at 12 months compared to standard medical therapy. |
| 80 | Surder et. Al. 2013 | Switzerland | Bone Marrow | BMMNCs | 200 | 166 | 34 | BMMNCs | 128 | Control | 67 | 56 | IC | 0.5 | 7 | 30 | 4 | High | Intracoronary infusion of bone marrow-derived mononuclear cells (BM-MNC) at either 5–7 days or 3–4 weeks after acute myocardial infarction (AMI) did **not** improve left ventricular function at 4-month follow-up compared to standard medical therapy. |
| 81 | Nair et. Al. 2015 | India | Bone Marrow | BMMNCs | 250 | 220 | 30 | BMMNCs | 125 | Control | 125 | 48.07 | IC | 50 | 0.2 | 5 | 6 | High | Intracoronary infusion of autologous bone marrow-derived mononuclear cells (MNCs) did not significantly improve left ventricular ejection fraction (LVEF) at 6 months post-AMI compared to standard therapy, though a higher cell dose (≥5 × 10^8) showed a non-significant trend toward benefit. |
| 82 | Tendera et. Al. 2009 | Poland | Bone Marrow | BMMNCs | 200 | 137 | 63 | BMMNCs | 160 | Control | 40 | 59 | IC | 17.8 | 3 | 7 | 6 | High | Intracoronary infusion of selected (CD34+CXCR4+) or non-selected bone marrow cells did not significantly improve LVEF at 6 months in STEMI patients with reduced ejection fraction, though a trend was observed in those with severe baseline LV dysfunction. |
| 83 | Traverse et. Al. 2018 | USA | Bone Marrow | BMMNCs | 120 | 109 | 11 | BMMNCs | 79 | Control | 41 | 56 | IC | 15 | 3 | 7 | 24 | High | Bone marrow mononuclear cell (BMC) therapy did not improve left ventricular function recovery over 2 years in STEMI patients compared to placebo, and microvascular obstruction (MVO) was associated with worse outcomes. |
| 84 | Turan et. Al. 2012 | Germany | Bone Marrow | BMMNCs | 62 | 42 | 20 | BMMNCs | 42 | Control | 20 | 61 | IC | 1.5 | 7 | 7 | 12 | High | Intracoronary transplantation of freshly isolated bone marrow cells (BMCs) in acute myocardial infarction (AMI) patients significantly improved left ventricular ejection fraction (LVEF), reduced infarct size, and enhanced long-term mobilization of bone marrow-derived progenitor cells (BM-CPCs) compared to controls. |
| 85 | Laan et. Al. 2011 | Netherlands | Bone Marrow | BMMNCs | 60 | 50 | 10 | BMMNCs | 41 | Control | 19 | 55 | IC | 29.1 | 1 | 7 | 4 | High | Adjuvant therapy with BMMCs or PBMCs did not improve microcirculation recovery in STEMI patients after primary PCI, refuting the hypothesis of enhanced neovascularization. |
| 86 | Wang et. Al. 2014 | China | Bone Marrow | BMMSCs | 58 | 35 | 23 | BMMSCs | 28 | Control | 30 | 58 | IC | 14 | 8 | 1 | 6 | High | Intracoronary autologous BMSC transplantation reduced early adverse events and rehospitalization rates in AMI patients post-PCI but did not significantly improve LVEF or infarct size compared to controls. |
| 87 | Wollert et. Al. 2004 | Germany | Bone Marrow | BMMSCs | 60 | 42 | 18 | BMMSCs | 30 | Control | 30 | 59.2 | IC | 245 | 1 | 8 | 6 | High | Intracoronary transfer of autologous bone-marrow cells significantly improved left-ventricular ejection fraction (LVEF) by 6.7 percentage points at 6 months compared to controls (p=0.0026), with no increased risk of adverse events. |
| 88 | Yao et. Al. 2009 | China | Bone Marrow | BMMSCs | 39 | 33 | 6 | BMMSCs | 27 | Control | 12 | 52 | IC | 19 | 7 | 30 | 12 | High | Repeated intracoronary infusion of autologous bone marrow mononuclear cells significantly improved left ventricular function and reduced infarct size in patients with large acute myocardial infarction compared to single infusion or placebo. |
